# Type 2 Diabetes Sub-Phenotypes and Their Association with Cardiovascular Disease Risk: A Multi-Center Study

**DOI:** 10.1101/2025.03.09.25323601

**Authors:** Kan Wang, Raymond Noordam, Stella Trompet, Julie A.E. van Oortmerssen, J. Wouter Jukema, M. Kamran Ikram, Jana Nano, Christian Herder, Annette Peters, Christian Gieger, Barbara Thorand, Maryam Kavousi, Fariba Ahmadizar

**Author notes:** Corresponding author: Dr. Fariba Ahmadizar, PharmD, MSc, PhD.

## Abstract

**Aims/Hypothesis:** Type 2 diabetes mellitus (T2D) is a heterogeneous condition influenced by lipid metabolism, inflammation, and genetic predisposition, all of which contribute to variable cardiovascular disease (CVD) risk. Identifying robust T2D sub-phenotypes and understanding their interactions with genetic predisposition is critical for personalized CVD risk assessment and care. This study aims to derive clinically relevant T2D sub-phenotypes and assess their association with CVD risk by employing robust methodology and replication across cohorts.

**Methods:** We analyzed data from the Rotterdam Study (n=1,250), applying Gaussian mixture clustering to derive T2D sub-phenotypes based on nine metabolic risk factors: age at diabetes diagnosis, sex, body mass index (BMI), fasting blood glucose, HOMA-IR, cholesterol levels (total, HDL, LDL), and C-reactive protein (CRP). Cox proportional hazard models adjusted for confounders were used to estimate hazard ratios (HRs) and 95% confidence intervals (CIs) for associations between T2D sub-phenotypes and a composite CVD outcome (coronary heart disease and stroke). Kaplan-Meier (KM) survival curves were created to study the risk of incident CVD across T2D sub-phenotypes, with the lowest-risk sub-phenotype as the reference group. Polygenic risk scores (PRS) for T2D, divided into tertiles, were included to explore the interaction of genetic predisposition with diabetes sub-phenotypes. Clustering was replicated in the KORA (n=243) and PROSPER (n=179) cohorts, with association analyses validated in the KORA cohort. We considered effect size and confidence intervals, not just p-values, for comprehensive result interpretation.

**Results:** Three distinct T2D sub-phenotypes emerged: (1) an “unspecified” sub-phenotype (53.4%) with lower levels of metabolic risk factors, (2) an “insulin-resistant” sub-phenotype (23.8%) characterized by higher BMI, HOMA-IR, and CRP, and (3) a “dyslipidemic” sub-phenotype (22.3%) with elevated total and LDL-cholesterol. Compared to the dyslipidemic sub-phenotype (reference group based on KM analyses), the adjusted HR for incident CVD was 1.04 (95% CI: 0.76, 1.42) for the unspecified sub-phenotype and 1.20 (95% CI: 0.84, 1.72) for the insulin-resistant sub-phenotype, indicating a slightly elevated risk of CVD for the insulin-resistant sub-phenotype. Among individuals with high T2D PRS, the insulin-resistant sub-phenotype exhibited the highest CVD risk (HR 2.28, 95% CI 1.13, 4.60) compared to low and medium PRS from T2D. The robustness of the sub-phenotypes and their associations with CVD risk was confirmed in independent KORA and PROSPER cohorts.

**Conclusions/Interpretation:** These findings emphasize the importance of understanding metabolic and clinical diversity within T2D to better guide personalized management strategies. Further research through longitudinal studies, diverse populations, and advanced molecular profiling is essential to refine sub-phenotypic classifications and uncover underlying mechanisms to enhance patient outcomes

## Introduction

Type 2 diabetes mellitus (T2D), conventionally diagnosed based on hyperglycemia, is a multifaceted and heterogeneous condition influenced by various metabolic, inflammatory, and genetic factors [1, 2]. Complications of T2D, especially cardiovascular disease (CVD), stem from complex interactions among these factors. This complexity, coupled with conventional classification methods and a lack of personalized management, may contribute to suboptimal control of diabetes-related complications [3, 4]. The spectrum model of diabetes highlights that T2D arises from dysfunction across multiple etiological pathways, including beta cell function, insulin action, glucagon regulation, and fat distribution. Although each T2D case may manifest a unique combination of these defects, clinical practice often overlooks this heterogeneity [5].

Data-driven cluster analysis offers a promising method for addressing T2D heterogeneity [6, 7]. Ahlqvist et al. showed the potential of clustering to identify distinct T2D subgroups based on factors like autoantibodies, age, BMI, HbA1c, β-cell function, and insulin resistance [8]. Subsequent studies have explored the clinical utility of these clusters, including our own work in the Chinese population [9–11]. However, previous clustering approaches did not account for lipid and inflammatory biomarkers, which are known to play essential roles in metabolic syndrome and insulin resistance [12].

In this study, we leveraged data from the Rotterdam Study to identify distinct T2D sub-phenotypes based on metabolic and inflammatory markers and to examine their association with future CVD risk. We also assessed whether genetic predisposition, quantified by PRS, modified the relationship between T2D sub-phenotypes and CVD. We hypothesize that individuals with higher PRS for T2D will show stronger associations between specific T2D sub-phenotypes and CVD risk, providing insight into how genetic factors modify these relationships [13]. We further validated our findings by replicating these clusters in the KORA and PROSPER cohorts and examining their associations with CVD outcomes in the KORA cohort. This approach aims to enhance our understanding of T2D heterogeneity and inform personalized management strategies tailored to specific phenotypic profiles.

## Method

### Study setting

We used three databases for this analysis: the Rotterdam study as the derivation cohort and the KORA and PROSPER studies as the validation cohorts.

**The Rotterdam study** is a prospective cohort study of community-dwelling adults aged 55 and older in Rotterdam, the Netherlands, with a detailed study design reported elsewhere [14]. Briefly, the baseline examination for the first cohort was completed between 1990 and 1993 (RS-I) with 7,983 participants aged 55 years or over. The study was extended in 2000, with the second cohort of individuals who had reached 55 years or moved into the study area after 1990 (RS-II, n = 3011). In 2006, a third cohort was enrolled, including inhabitants aged 45 years and older (RS-III, n = 3932). In 2016, the most recent extension of the cohort was set up with 3,005 persons aged 40 years and over. By the start of 2021, the Rotterdam Study comprised 17,931 participants aged 40 years or over, with a response rate for all four cycles at a study entry of 65% (17,931 out of 27,571). The Rotterdam Study has been approved by the Medical Ethics Committee of the Erasmus MC (registration number MEC 02.1015) and by the Dutch Ministry of Health, Welfare and Sport (Population Screening Act WBO, license number 1071272-159521-PG). The Rotterdam Study Personal Registration Data collection is filed with the Erasmus MC Data Protection Officer under registration number EMC1712001. The Rotterdam Study has been entered into the Netherlands National Trial Register (NTR; https://onderzoekmetmensen.nl/en/trial/23050) and the WHO International Clinical Trials Registry Platform (ICTRP; https://www.who.int/clinical-trials-registry-platform, search portal https://trialsearch.who.int/) under shared catalogue number NTR6831. Almost all (98%) participants provided written informed consent to participate in the study and obtain their information from treating physicians.

**The KORA study** consists of three World Health Organization (WHO) MONICA (Monitoring of Trends and Determinants in Cardiovascular Disease) studies (S1, S2, and S3) that were started during the 1980s to 1990s as a general effort of WHO to generate population-based epidemiological data in several areas in Europe [15]. The MONICA Augsburg studies S1-S3 were then carried on and further developed within the frame of KORA with the addition of a new cohort (S4) in 1999-2001 and several follow-up examinations as well as regular questionnaire-based follow-up waves and mortality follow-up [15]. The present analysis includes all participants with T2D from the KORA F4 study (2006-2008; total n=3080), the first follow-up examination of KORA S4 [16]. All study participants provided written informed consent, and the Bavarian Medical Association ethics committee has approved the KORA studies.

**The PROSPER study** The PROspective Study of Pravastatin in the Elderly at Risk (PROSPER) is a prospective multicenter randomized placebo-controlled trial designed to evaluate the potential of pravastatin in reducing major cardiovascular events among elderly individuals [17, 18]. Conducted between December 1997 and May 1999, the trial enrolled 5,804 participants aged between 70 and 82 years from Scotland, Ireland, and the Netherlands. Participants were selected based on their preexisting vascular disease or increased risk factors such as a history of smoking, hypertension, or diabetes. Ethical approval was obtained from the institutional review boards of all participating centers, and written informed consent was obtained from all participants.

### Inclusion and Exclusion Criteria

The study population included all prevalent and newly identified cases of T2D. In the Rotterdam Study, T2D cases were identified according to fasting glucose >6.9 mmol/L, non-fasting glucose >11.0 mmol/L, or glucose-lowering medications.

In KORA, prevalent clinically diagnosed T2D was initially defined based on a clinical T2D diagnosis or self-reported use of glucose-lowering medications. Subsequently, this information was confirmed by medical records or by contacting the treating physicians [16]. Newly identified T2D cases comprised participants with fasting glucose concentrations and/or 2 h glucose concentrations after an oral glucose tolerance test (OGTT) in the diabetic range according to the 1999 WHO criteria [19]. In PROSPER, T2D was defined based on diagnosis or fasting blood glucose > 7mmol/L.

For the longitudinal study, individuals with prevalent CVD, defined as coronary heart disease (CHD) and stroke, were excluded.

### Measurements

***Cluster variables*** included the age at T2D diagnosis (in years), sex (male or female), body mass index (BMI) (in kg/m^2^), fasting blood glucose (FBG) (in mmol/L), HOMA-IR index (calculated as the product of fasting insulin (mU/l) and FBG (mmol/l), divided by 22.5, serves as a robust marker of insulin resistance), total cholesterol (in mg/dL), high-density lipoprotein (HDL)-cholesterol (in mg/dL), low-density lipoprotein (LDL)-cholesterol (in mg/dL), and C-reactive protein (CRP) (in mg/L and log-transformed). We adjusted the total cholesterol values for individuals taking lipid-lowering medication by dividing their values by 0.8; no adjustment was made to HDL-cholesterol [20]. LDL-cholesterol was calculated with the updated equation for subjects with normolipidemia and/or hypertriglyceridemia, with modified total cholesterol used in this formula [21]. If the measured LDL-cholesterol was available, we divided its value by 0.7 for individuals on lipid-lowering medication [20]. All these cluster variables are well-captured in routine clinical care and reflect a range of measures known to be altered in T2D and associated with variation in risk of adverse diabetes outcomes to investigate potential T2D sub-phenotypes.

All these cluster variables were measured simultaneously as diabetes or if data was unavailable within three years before or one year after T2D diagnosis. If multiple recordings were available, the closest recording to the T2D diagnosis date was used in the analysis.

#### Ascertainment of incident cardiovascular disease

According to the International Classification of Disease 9^th^ or 10^th^ Revisions (ICD 9 or ICD 10), incident CVD was defined as a composite endpoint including CHD and stroke occurring during the follow-up in persons without a previous CHD or stroke at baseline. In the Rotterdam Study, these clinical outcomes were collected continuously through an automated follow-up system involving digital linkage of the study database to medical records maintained by general practitioners working in the research area. Also, information on vital status is obtained from the central registry of the municipality of the city of Rotterdam. CHD was defined as myocardial infarction (MI) and surgical or percutaneous coronary revascularization procedure (as a proxy for unstable or incapacitating angina). MI was defined according to the triad of symptoms, indicative electrocardiogram changes, and cardiac biomarkers. Stroke was described as a syndrome of rapidly developing clinical signs of focal (or global) disturbance of cerebral function. Follow-up extended from baseline (1997) to the end of (2015) [22]. In the KORA study, CVD was defined as a combined endpoint of incident CHD or stroke. Incident CHD comprised non-fatal MI, coronary death, and sudden death. Incident stroke comprised non-fatal and fatal strokes without transient ischemic attacks. Follow-up extended from baseline (2006 - 2008) to the end of 2016. Further information on incident CHD and stroke assessment procedures in KORA can be found elsewhere [23, 24]. The PROSPER cohort had a shorter follow-up period, limiting its ability to assess longitudinal outcomes.

#### Covariates

Confounders were included to account for potential sources of bias that could distort the relationship between cluster variables and cardiovascular outcomes. These covariates included age, sex, education level classified into primary education, lower/intermediate general education, vocational/higher general education, and higher vocational/university, smoking status categorized as former, current, or never smoker, systolic blood pressure, and the use of blood pressure-lowering medication.

#### Ascertainment of diabetes-related polygenic risk score

##### Genotyping

In the Rotterdam Study, genotyping has been performed using the Illumina 550K and 610K quad array (Illumina Inc., San Diego, CA, USA) imputed to the Haplotype Reference Consortium reference panel (version 1.0) with Minimac 3. We included independent genetic variants associated with T2D based on a GWAS on individuals of European ancestry to calculate a weighted PRS [25]. In the KORA study, genotyping and calling were performed using the Affymetrix Axiom platform and software. Imputation was performed using minimac3 (Michigan Imputation Server) and the reference panel Haplotype Reference Consortium (Panel (r1.1), April 2016). In the PROSPER study, genotyping was performed with the Illumina 660K bead chip; after QC (call rate <95%), 5,244 subjects and 557,192 SNPs were left for analysis. These SNPs were imputed to 2.5 million SNPs based on the HAPMAP built 36 with MACH imputation software and in the next step to the Haplotype Reference Consortium panel (version 1.0).

Genetic risk was evaluated using a polygenic risk score (PRS) for T2D calculated as the sum of the products of the single-nucleotide polymorphism (SNP) allele dosages of the 403 genetic variants and their respective reported effect estimates [25].

##### Statistical analyses

###### Data preparation

We identified both prevalent (at the enrollment of sub-cohorts) and newly identified (during follow-up) T2D cases. In a complete case analysis approach, all nine cluster variables were firstly adjusted by the age of T2D diagnosis and sex by taking the residuals of a linear model with the respective biomarker as an outcome. The residuals were centered and standardized, and those with extreme outliers >5 SDs from the mean were excluded. All downstream analyses were stratified by case status (prevalent or newly identified cases) to check for possible deviations caused by treatment and to assess the stability of observed phenotypes during the natural course of the disease.

###### Cluster analysis

We used Gaussian mixture modelling (GMM), a probabilistic unsupervised learning technique, with a diagonal covariance matrix to identify clusters separately for prevalent and newly identified T2D cases. It is trained using the Expectation-Maximization (EM) algorithm to optimize parameters. While GMM is designed for continuous variables, categorical data can be handled through latent class analysis, which models categorical data as mixtures of discrete latent classes [26].

We applied three metrics to determine the optimal number of clusters: Bayesian Information Criterion (BIC), average silhouette width, and the gap statistic. BIC prioritizes model fit while penalizing complexity, with lower values indicating better fit; the silhouette width measures clustering quality by comparing within-cluster cohesion to separation from other clusters; and the gap statistic assesses how structured the clustering is compared to a random reference distribution. After independently evaluating each metric, we compared the results to select the final number of clusters. The final decision on the number of clusters was based on combining these methods. We analyzed baseline characteristics by each T2D sub-phenotype in the Rotterdam Study. We first computed descriptive statistics, including means and SD for continuous variables and frequencies for categorical variables across the T2D sub-phenotype. We used ANOVA for continuous variables and Chi-Square tests for categorical variables to compare cluster characteristics.

##### Survival analysis

Kaplan-Meier (KM) survival curves were generated to illustrate the risk of incident CVD across T2D sub-phenotypes, with the sub-phenotype exhibiting the lowest risk designated as the reference group. Cox proportional hazard regression models were employed to examine the association between T2D sub-phenotypes and incident CVD during follow-up in the Rotterdam Study. Model 1 was adjusted for age at baseline and sex, while model 2 included additional adjustments for education, smoking status, systolic blood pressure, and use of blood pressure-lowering medication. Further analyses stratified these associations by tertiles of the PRS for T2D.

##### External Replication

An external replication of the cluster analysis was performed using data from the KORA and PROSPER studies. The same clustering method and nine cluster variables employed in the Rotterdam Study were applied independently to these cohorts, identifying the three T2D sub-phenotypes with similar patterns. Due to the limited sample size, the survival analysis replicated in the KORA study only included model 1 (adjusted for age and sex).

In the PROSPER Study, follow-up data were limited to less than four years, reducing the ability to assess long-term CVD outcomes robustly. Therefore, data from the PROSPER Study were included as external validation of only the clustering approach.

All analyses were performed in R (version 4.4.2).

## Results

### Clustering, discovery and replication

GMM analysis was performed in the Rotterdam Study to identify sub-phenotypes of T2D. The optimal number of clusters was determined based on several criteria: the BIC value (**ESM Figure 1**), the average silhouette width (**ESM Figure 2**), and the gap statistic (**ESM Figure 3**). These analyses revealed three distinct T2D sub-phenotypes, initially identified separately for prevalent and newly identified T2D cases (**Figure 1**). The pattern and distribution of cluster variables were similar between the two groups, indicating that they could be analyzed together. To increase the sample size for the cluster analysis, the related clusters from both groups were combined, delineating three distinct T2D sub-phenotypes: “dyslipidemic”, “unspecified”, and “insulin-resistant”.

**Figure 1.**
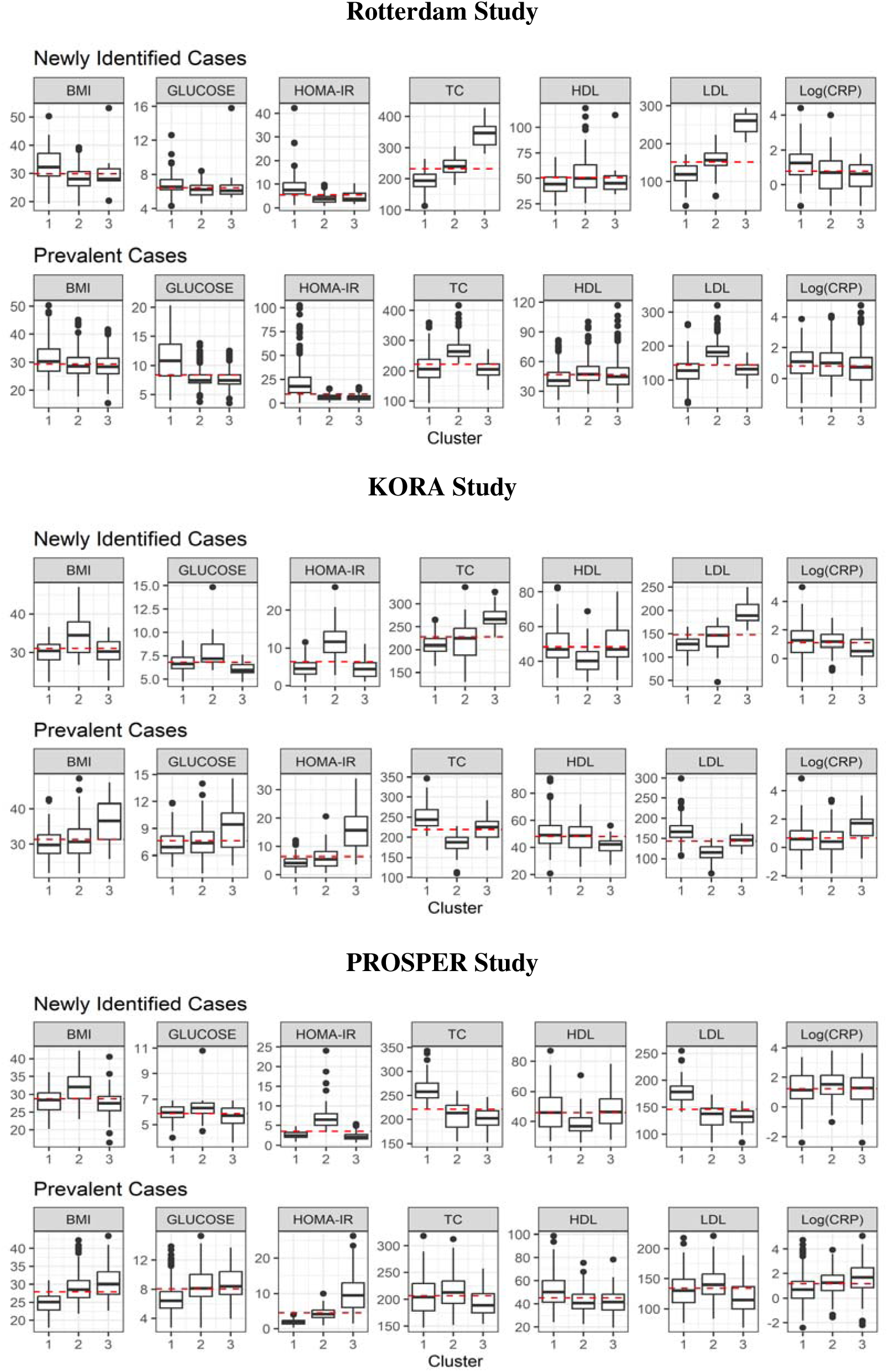
Distribution of cluster variables among different type 2 diabetes sub-phenotypes across the three databases. **Note.** Given that the number of optimal clusters and the distribution of cluster variables were similar among newly identified and prevalent cases in all cohorts, the identical phenotypes were further combined as *unspecified sub-phenotypes* (cluster 2 for newly identified cases and cluster 3 for prevalent cases in the Rotterdam Study, cluster 1 for newly identified cases and cluster 2 for prevalent cases in the KORA Study, cluster 3 for newly identified cases and cluster 1 for prevalent cases in the PROSPER Study); *insulin-resistant sub-phenotypes* (cluster 1 for newly identified cases and cluster 1 for prevalent cases in the Rotterdam Study, cluster 2 for newly identified cases and cluster 3 for prevalent cases in the KORA Study, cluster 2 for newly identified cases and cluster 3 for prevalent cases in the PROSPER Study); and *dyslipidemic sub-phenotypes* (cluster 3 for newly identified cases and cluster 2 for prevalent cases in the Rotterdam Study, cluster 3 for newly identified cases and cluster 1 for prevalent cases in the KORA Study, cluster 1 for newly identified cases and cluster 2 for prevalent cases in the PROSPER Study).

Using data from the KORA and PROSPER studies and based on the derived curves of the BIC value, average silhouette width, and gap statistic (**ESM Figure 1-3)**, three distinct T2D sub-phenotypes were identified. These sub-phenotypes exhibited metabolic profiles similar to those observed in the Rotterdam Study (**Table 1, ESM Table 1&2**).

**Table 1.** Characteristics of the included participants for different type 2 diabetes phenotypes in the Rotterdam Study.

| **T2D Sub-phenotypes** | **Dyslipidemic sub-phenotype** | **Unspecified sub-phenotype** | **Insulin-resistant sub-phenotype** | ***P*-value** |
| --- | --- | --- | --- | --- |
| No. of participants | 286 | 667 | 297 |  |
| Diabetes status, n (%) |  |  |  | <0.001 |
| Prevalent cases | 276 (96.5) | 534 (80.1) | 230 (77.4) |  |
| Newly identified cases | 10 (3.5) | 133 (19.9) | 67 (22.6) |  |
| Age, years, mean (SD) | 66.8 (8.5) | 67.2 (9.6) | 66.5 (9.3) | 0.512 |
| Sex, male, n (%) | 142 (49.7) | 331 (49.6) | 153 (51.5) | 0.852 |
| Education, n (%) |  |  |  | 0.548 |
| Primary education | 38 (13.4) | 110 (16.6) | 47 (15.9) |  |
| Lower/intermediate general education | 121 (42.6) | 275 (41.4) | 125 (42.2) |  |
| Vocational/higher general education | 90 (31.7) | 175 (26.4) | 82 (27.7) |  |
| Higher vocational/university | 35 (12.3) | 104 (15.7) | 42 (14.2) |  |
| Current smoking, n (%) | 55 (19.6) | 153 (23.4) | 59 (20.3) |  |
| Systolic blood pressure, mmHg, mean (SD) | 147.9 (23.7) | 146.6 (22.0) | 146.7 (21.3) | 0.725 |
| Use of blood pressure-lowering medication, n (%) | 143 (51.3) | 323 (48.9) | 169 (59.1) | 0.015 |
| Body mass index, kg/m<sup>2</sup>, mean (SD) | 29.4 (4.8) | 28.7 (4.0) | 31.5 (5.8) | <0.001 |
| Fasting blood glucose, mmol/L, mean (SD) | 7.7 (1.7) | 7.3 (1.5) | 10.2 (3.6) | <0.001 |
| HOMA-IR*, median (IQR) | 5.8 (3.9, 8.0) | 5.0 (3.3, 7.3) | 14.6 (8.4, 22.6) | <0.001 |
| Total cholesterol, mg/L, mean (SD) | 272.2 (34.7) | 211.7 (29.0) | 205.4 (43.7) | <0.001 |
| HDL-cholesterol, mg/L, mean (SD) | 49.1 (12.0) | 48.5 (15.1) | 43.6 (12.1) | <0.001 |
| LDL-cholesterol, mg/L, mean (SD) | 189.1 (28.8) | 135.4 (24.5) | 125.8 (35.4) | <0.001 |
| Use of lipid-lowering medication, n (%) | 96 (33.6) | 154 (23.1) | 77 (25.9) |  |
| CRP, mg/L, median (IQR) | 2.7 (1.1, 5.2) | 2.0 (0.8, 3.7) | 3.0 (1.4, 5.5) | <0.001 |
| Polygenic risk score, median (IQR) | 27.1 (26.5, 27.6) | 27.0 (26.5, 27.5) | 27.0 (26.6, 27.5) | 0.456 |
| Tertile of polygenic risk score, n (%) |  |  |  | 0.259 |
| Lower | 82 (32.7) | 197 (35.0) | 77 (30.3) |  |
| Medium | 74 (29.5) | 189 (33.6) | 93 (36.6) |  |
| High | 95 (37.8) | 177 (31.4) | 84 (33.1) |  |

**Abbreviations:** SD, standard deviation; HOMA-IR, Homeostatic Model Assessment of Insulin Resistance; IQR, interquartile range; HDL, high-density lipoprotein; LDL, low-density lipoprotein; CRP, c-reactive protein.
\* **HOMA-IR** index calculated as fasting insulin (mU/l) × FBS (mmol/l)/22.5

### Baseline Characteristics

Baseline characteristics for participants in the Rotterdam Study are detailed in **Table 1 & Figure 1**. 22.9% were classified within the dyslipidemic T2D sub-phenotype, primarily distinguished by elevated total cholesterol and LDL-cholesterol levels. The unspecified sub-phenotype accounted for a notable proportion of 53.4% of participants, with lower metabolic risk factors than the dyslipidemic and insulin-resistant sub-phenotypes. For instance, the mean (SD) BMI was 28.4 (4.0) compared to 29.4 (4.8) and 31.5 (5.8), mean fasting blood glucose was 7.3 (1.5) compared to 7.7 (1.7) and 10.2 (3.6), and median HOMA-IR was 5.0 (3.3, 7.3) compared to 5.8 (3.9, 8.0) and 14.6 (8.4, 22.6), respectively.

The insulin-resistant sub-phenotype, comprising 23.8% of T2D cases, showed elevated BMI, FBG, HOMA-IR, and CRP levels.

Across the three sub-phenotypes, CVD-related factors, such as smoking status and systolic blood pressure, were relatively consistent.

Additionally, we examined the distribution of PRS for T2D across these sub-phenotypes and observed no significant differences (p = 0.46).

The KORA (**ESM Table 1)** and PROSPER (**ESM Table 2)** observed a similar distribution of sub-phenotypes and corresponding CVD risk profiles, further supporting the consistency of these findings across different cohorts.

**Table 2.** Risk of incident cardiovascular disease among different type 2 diabetes sub-phenotypes.

| Cohorts | T2D Sub-phenotypes | Participants | Person-years at risk | Events |  | HR (95%CI) |
| --- | --- | --- | --- | --- | --- | --- |
| **Rotterdam Study** | Dyslipidemic | 286 | 2684 | 62 |  | 1.00 (ref) |
|  | Unspecified | 667 | 6023 | 151 | Model 1 | 1.01 (0.75, 1.36) |
|  |  |  |  |  | Model 2 | 1.04 (0.76, 1.42) |
|  | Insulin-resistant | 297 | 2517 | 71 | Model 1 | 1.20 (0.85, 1.69) |
|  |  |  |  |  | Model 2 | 1.20 (0.84, 1.72) |
| **KORA Study**<sup>#</sup> | Dyslipidemic | 98 | 774 | 13 | Model 1 | 1.00 (ref) |
|  | Unspecified | 110 | 815 | 17 | Model 1 | 1.22 (0.59, 2.51) |
|  | Insulin-resistant | 35 | 241 | 6 | Model 1 | 1.45 (0.55, 3.83) |

**Abbreviations:** HR, hazard ratio; CI, confidence interval
Model 1: Adjusted for age, sex
Model 2: Model 1 plus education, smoking, systolic blood pressure, and use of blood pressure-lowering medication.
<sup>#</sup> Model 2 couldn't be implemented due to the sample size limit.

### Diabetes Sub-phenotypes, Genetic Risk, and Incident Cardiovascular Disease

Following the recommendations of the American Statistical Association, we avoided relying solely on p-values and statistical significance [27]. Instead, we also considered effect size and precision (confidence intervals) to interpret our results more comprehensively.

KM analyses indicated that the T2D dyslipidemic sub-phenotype had the lowest CVD risk, serving as the reference group in the subsequent Cox regression analyses (**ESM Figure 4)**.

#### Rotterdam Study

Among 1,250 participants followed over 11,224 person-years, 284 incident CVD events were recorded. For prevalent T2D, the start date was the Rotterdam Study visit date; for newly identified T2D, the start date was the date of diabetes diagnosis. Median follow-up time was 8.15 years [IQR: 5.95-13.67] overall, with 8.32 years [IQR: 6.43–13.90] for prevalent cases and 7.23 years [IQR: 5.17–11.05] for newly identified cases. Baseline cluster membership identified 286 participants (2,684 person-years, 62 CVD events, incidence rate 23.1 per 1000 person-years) in the dyslipidemic sub-phenotype, 667 participants (6,023 person-years, 151 CVD events, incidence rate 25.1 per 1000 person-years) in the unspecified sub-phenotype, and 297 participants (2,517 person-years, 71 CVD events, incidence rate 28.2 per 1000 person-years) in the insulin-resistant sub-phenotype. In Model 1 (adjusted for age and sex), HRs for incident CVD were 1.01 (95% CI: 0.75–1.36) for the unspecified sub-phenotype and 1.20 (95% CI: 0.85–1.69) for the insulin-resistant sub-phenotype, with the dyslipidemic sub-phenotype as the reference. Model 2 (further adjusted for education, smoking, systolic blood pressure, and use of blood pressure-lowering medication) yielded consistent HRs of 1.04 (95% CI: 0.76–1.42) and 1.20 (95% CI: 0.84–1.72), respectively (**Table 2**).

**Table 3** presents the risk of incident CVD across different T2D sub-phenotypes stratified by PRS tertiles for T2D. The results highlight variations in HR for CVD events among low, medium, and high PRS groups for each sub-phenotype. Both models 1 and 2 showed consistent trends, with the increased risk particularly evident for the insulin-resistant sub-phenotype in the highest PRS tertile (2.28, 95% CI: 1.13–4.60).

**Table 3.** Risk of incident cardiovascular disease among different type 2 diabetes sub-phenotypes, by polygenic risk score tertiles.

| Cohorts |  | PRS | Dyslipidemic sub-phenotype |  | Unspecified sub-phenotype |  | Insulin-resistant sub-phenotype |  |
| --- | --- | --- | --- | --- | --- | --- | --- | --- |
|  |  |  | Num. of events<br>/ Participants | HR (95%CI) | Num. of events<br>/ Participants | HR (95%CI) | Num. of events<br>/ Participants | HR (95%CI) |
| **Rotterdam**<br>**Study \*** | Model 1 | Low | 21 / 82 | 1.00 (ref) | 51 / 197 | 1.01 (0.60, 1.68) | 17 / 77 | 0.94 (0.49, 1.78) |
|  |  | Medium | 15 / 74 | 1.00 (ref) | 35 / 189 | 1.08 (0.59, 1.99) | 20 / 93 | 1.22 (0.62, 2.38) |
|  |  | High | 16 / 95 | 1.00 (ref) | 40 / 177 | 1.02 (0.56, 1.84) | 23 / 84 | 1.73 (0.91, 3.31) |
|  | Model 2 | Low | 21 / 82 | 1.00 (ref) | 51 / 197 | 1.03 (0.60, 1.74) | 17 / 77 | 0.83 (0.42, 1.62) |
|  |  | Medium | 15 / 74 | 1.00 (ref) | 35 / 189 | 0.97 (0.50, 1.86) | 20 / 93 | 1.12 (0.55, 2.30) |
|  |  | High | 16 / 95 | 1.00 (ref) | 40 / 177 | 1.49 (0.78, 2.83) | 23 / 84 | **2.28 (1.13, 4.60)** |
| **KORA**<br>**Study #** | Model 1 | Low | 1 / 30 | 1.00 (ref) | 5 / 24 | 6.44 (0.75, 55.58) | 1 / 15 | 1.78 (0.11, 29.40) |
|  |  | Medium | 3 / 24 | 1.00 (ref) | 6 / 37 | 0.92 (0.21, 3.97) | 1 / 8 | 0.71 (0.07, 7.31) |
|  |  | High | 7 / 32 | 1.00 (ref) | 5 / 31 | 0.82 (0.26, 2.61) | 3 / 6 | **4.22 (1.06, 16.71)** |

**Abbreviations:** HR, hazard ratio; CI, confidence interval; PRS, polygenic risk score
\* Model 1: Adjusted for age, sex,
Model 2: Model 1 plus education, smoking, systolic blood pressure, and use of blood pressure-lowering medication.
The p value for interaction was 0.049.

#### KORA Study

In this validation cohort, 243 participants were followed over 1,830 person-years and 36 incident CVD events were recorded. The median total follow-up time was 8.40 [IQR: 6.98–8.94] years, with 8.44 years [IQR: 6.98–8.96] for prevalent cases and 8.33 years [IQR: 6.88–8.89] for newly identified cases. Baseline cluster membership included 98 participants (774 person-years, 13 events, incidence rate 16.8 per 1000 person-years) in the dyslipidemic sub-phenotype, 110 participants (815 person-years, 17 events, incidence rate 20.9 per 1000 person-years) in the unspecified sub-phenotype, and 35 participants (241 person-years, 6 events, incidence rate 24.9 per 1000 person-years) in the insulin-resistant sub-phenotype. Model 1 adjusted HRs were 1.22 (95% CI: 0.59–2.51) for the unspecified sub-phenotype and 1.45 (95% CI: 0.55–3.83) for the insulin-resistant sub-phenotype, with the dyslipidemic sub-phenotype as the reference. Model 2 could not be implemented due to sample size limitations (**Table 2).**

In PRS-stratified analyses, the KORA Study reflects similar patterns, with varying levels of risk across sub-phenotypes. The insulin-resistant T2D sub-phenotype in the high PRS tertile exhibited an elevated risk (HR: 4.22, 95% CI: 1.06–16.71) (**Table 3**).

## Discussion

Using a data-driven clustering approach in the Rotterdam Study, we identified three distinct sub-phenotypes of T2D: dyslipidemic (distinguished by high total cholesterol and LDL-cholesterol levels; 23% of T2D), unspecified (characterized by relatively lower levels of cluster-defining variables; 53% of T2D cases), and insulin-resistant (marked by elevated BMI, FBG, HOMA-IR, and CRP; 24% of T2D). These findings were independently validated in the KORA and PROSPER cohorts. The PROSPER cohort was only utilized to replicate the clustering analysis due to its shorter follow-up period, precluding longitudinal outcome assessment.

Each sub-phenotype exhibited unique metabolic and clinical profiles. The insulin-resistant sub-phenotype was associated with an increased risk of CVD, particularly among individuals with a high PRS for T2D, confirmed by the KORA Study. While these findings provide insights into the heterogeneity of T2D and its association with CVD risk, further research is needed to determine causality and evaluate the predictability of CVD risk based solely on sub-phenotypic classification.

The clustering underscores that not all T2D patients exhibit the same pathophysiological characteristics, which is essential for developing personalized therapeutic strategies [28]. Our findings raise important considerations regarding the clinical utility of clustering in T2D. Identifying the most clinically meaningful clusters, prioritizing variables that effectively capture metabolic heterogeneity, and understanding their implications for risk stratification, early intervention, and personalized treatment strategies are crucial for advancing patient care [29].

Interestingly, the dyslipidemic sub-phenotype, comprising 23% of T2D cases, appeared protective against CVD events according to the KM analyses, which may seem counterintuitive given the general association between dyslipidemia and increased CVD risk. However, this observation could be explained by several factors [30]. For example, this sub-phenotype may include individuals with a genetic makeup or lipid profile that confers a protective effect against CVD. While dyslipidemia impacts lipid transport and storage, its cardiovascular risk may also depend on insulin resistance, which directly drives vascular dysfunction through inflammation and endothelial damage [31]. This paradoxical effect may also stem from differences in the composition and function of lipid subtypes. The lipid composition within the dyslipidemic sub-phenotype, such as larger LDL particles, might explain its lower observed CVD risk [32]. Additionally, the balance between pro- and anti-atherogenic lipoproteins might influence CVD outcomes, with some lipid abnormalities being less harmful or even beneficial in certain metabolic contexts [32]. Genetic variations and environmental factors, including diet and lifestyle, could further modulate these effects. Future studies should explore advanced lipid profiling and gene-environment interactions to understand these relationships better.

The unspecified sub-phenotype, representing the majority (53%) of T2D cases in our study, was characterized by a less adverse risk factor profile based on the examined cluster variables, suggesting a less pronounced metabolic dysregulation than the insulin-resistant and dyslipidemic sub-phenotypes. This group likely reflects a heterogeneous population with milder or less specific metabolic disturbances, potentially representing an earlier or less advanced stage of T2D. The lower levels of CRP and lipid markers in this sub-phenotype may indicate a reduced immediate risk of complications, such as CVD, compared to the insulin-resistant group. Interestingly, there was no difference in the distribution of PRS for T2D across the sub-phenotypes, suggesting that genetic predisposition, as measured by PRS, does not significantly differ among the sub-phenotypes. This points to the potential importance of environmental and lifestyle factors and other non-genetic mechanisms in shaping the clinical presentation of T2D [33]. The heterogeneity within this sub-phenotype poses challenges for risk stratification and personalized management, as the absence of distinct metabolic features complicates targeted interventions. This group may also have unique genetic or environmental factors that buffer against severe metabolic derangements, warranting further exploration to uncover protective mechanisms. Longitudinal studies are needed to determine whether individuals in this group transition into more defined sub-phenotypes over time. Additional clinical and molecular variables, such as genetic, metabolomic, or inflammatory markers, could help refine this sub-phenotype, reduce its heterogeneity, and optimize individualized treatment strategies.

Our findings linking insulin resistance to increased CVD risk in T2D, though slightly, align with prior research [34–37]. Insulin resistance likely exacerbates CVD risk through multiple interrelated mechanisms, including systemic inflammation, endothelial dysfunction, and atherogenic dyslipidemia [35, 36, 38]. Individuals with insulin resistance often exhibit elevated levels of free fatty acids, a hallmark of this metabolic disturbance [39]. These fatty acids can damage blood vessels by promoting inflammation and interfering with the normal function of the endothelium—the thin layer of cells that lines blood vessels. This dysfunction can lead to the buildup of fatty deposits (atherosclerotic plaques) in the arteries [38]. Over time, these plaques can become unstable and rupture, causing blockages in the blood flow, which significantly increases the risk of cardiovascular events such as heart attacks or strokes. Stratified analyses suggested that individuals with high PRS for T2D within the insulin-resistant sub-phenotype had elevated CVD risk. This indicates that genetic predisposition modulates the relationship between metabolic disturbances and CVD outcomes. Integrating genetic risk scores into clinical practice could improve risk stratification and support personalized interventions, particularly for patients at high risk of complications [29].

This study’s strengths include its robust, data-driven clustering approach to classify T2D into distinct sub-phenotypes, reducing bias and enhancing validity. Validation in independent cohorts (KORA and PROSPER) supports the generalizability of the findings. Additionally, integrating genetic data, specifically PRS for T2D, provides valuable insights into the relationship between genetic predisposition and metabolic sub-phenotypes, broadening the understanding of disease heterogeneity. Despite these strengths, the study also has limitations that should be acknowledged. One main limitation is the cross-sectional nature of the cluster analysis, which provides a snapshot of the population at a single point in time. This limitation restricts our understanding of how individuals transition between sub-phenotypes over time and how these transitions influence the progression of complications like CVD. Future studies incorporating longitudinal measurements could elucidate the evolution of sub-phenotypes, improve risk stratification, and inform personalized disease management strategies [29]. Additionally, including both prevalent and newly identified T2D cases may introduce heterogeneity due to differences in disease duration and progression. For prevalent cases, the absence of exact diagnosis dates in the Rotterdam Study further constrained the calculation of precise follow-up periods for assessing CVD outcomes. Another limitation is the modest sample size, particularly for analyses stratified by genetic risk, which may have reduced the power to detect associations. The clustering approach prioritized established predictors of CVD risk, such as cholesterol subtypes, over potentially informative metrics like advanced lipid profiling or waist circumference. While clinically relevant, this focus may have limited the analysis’s granularity. Furthermore, the shorter follow-up period in the PROSPER cohort restricted its use to validation, precluding longitudinal outcome assessments. The predominantly European study population may also limit generalizability to other ethnic groups. Lastly, the absence of advanced molecular profiling, such as metabolomics or proteomics, left the underlying mechanisms of sub-phenotypic differentiation unexplored. Addressing these gaps with longitudinal designs, diverse cohorts, and multi-omics approaches will be essential to refining sub-phenotypic classifications and enhancing their clinical utility.

## Conclusion

This study provides important insights into the heterogeneity of T2D by identifying three distinct sub-phenotypes—dyslipidemic, unspecified and insulin-resistant—using a robust, data-driven clustering approach. The independent validation in two cohorts, KORA and PROSPER, underscores the generalizability and robustness of these findings. These sub-phenotypes not only highlight the diverse metabolic and clinical profiles within T2D but also suggest potential pathways linking specific sub-phenotypes, particularly the insulin-resistant group, to elevated CVD risk. While this work represents a significant step forward in understanding the complexity of T2D, it also emphasizes the need for longitudinal studies, diverse populations, and advanced molecular profiling to refine sub-phenotypic classifications further and explore their mechanistic underpinnings.

## Data Availability

All data produced in the present study are available upon reasonable request to the authors

## Acknowledgement

We thank all participants for their long-term commitment to the Rotterdam Study, KORA and PROSPER studies, the staff for data collection and research data management and the members who are responsible for the design and conduct of these studies.

