## Supplemental Table for "Type 2 Diabetes Sub-Phenotypes and Their Association with Cardiovascular Disease Risk: A Multi-Center Study"

**Supplement materials**

**ESM Table 1**. Characteristics of the included participants for different type 2 diabetes sub-phenotypes in the KORA Study

| **T2D Sub-phenotypes** | | **Dyslipidemic**  **sub-phenotype** | **Unspecified**  **sub-phenotype** | **Insulin-resistant**  **sub-phenotype** | ***P-value*** |
| --- | --- | --- | --- | --- | --- |
| No. of participants | | 98 | 110 | 35 |  |
| Diabetes status, n (%) | |  |  |  | <0.001 |
|  | Prevalent cases | 76 (77.6) | 66 (60.0) | 14 (40.0) |  |
|  | Newly identified cases | 22 (22.4) | 44 (40.0) | 21 (60.0) |  |
| Age, years, mean (SD) | | 66.5 (9.0) | 65.7 (9.5) | 65.7 (9.5) | 0.858 |
| Sex, male, n (%) | | 49 (50.0) | 67 (60.9) | 67 (60.9) | 0.283 |
| Body mass index, kg/m^2^, mean (SD) | | 30.3 (4.2) | 30.5 (4.4) | 30.5 (4.4) | <0.001 |
| Ever smoked, n (%) | | 52 (53.1) | 72 (65.5) | 72 (65.5) | 0.113 |
| Fasting blood glucose, mmol/L, mean (SD) | | 7.1 (1.4) | 7.2 (1.6) | 7.2 (1.6) | 0.01 |
| HOMA-IR^*^, median (IQR) | | 4.3 (2.9, 5.9) | 4.5 (3.1, 7.0) | 12.0 (9.1, 15.7) | <0.001 |
| Total cholesterol, mg/L, mean (SD) | | 254.5 (29.5) | 195.3 (25.0) | 224.5 (41.7) | <0.001 |
| HDL-cholesterol, mg/L, mean (SD) | | 50.0 (11.0) | 49.3 (11.0) | 41.9 (9.2) | 0.001 |
| LDL-cholesterol, mg/L, mean (SD) | | 173.8 (27.3) | 118.3 (19.5) | 144.8 (28.7) | <0.001 |
| Use of lipid-lowering medication, n (%) | | 40 (40.8) | 18 (16.4) | 7 (20.0) | <0.001 |
| CRP, mg/L, median (IQR) | | 1.7 (0.9, 3.2) | 2.3 (0.9, 4.9) | 3.2 (2.1, 6.0) | 0.008 |
| Polygenic risk score, median (IQR) | | 27.0 (26.5, 27.6) | 27.0 (26.6, 27.6) | 26.6 (26.1, 27.3) | 0.049 |
| Tertile of polygenic risk score, n (%) | |  |  |  | 0.072 |
|  | Lower | 30 (34.9) | 24 (26.1) | 15 (51.7) |  |
|  | Medium | 24 (27.9) | 37 (40.2) | 8 (27.6) |  |
|  | High | 32 (37.2) | 31 (33.7) | 6 (20.7) |  |

**Abbreviations:** SD, standard deviation; IQR, interquartile range; HOMA-IR, Homeostatic Model Assessment of Insulin Resistance; HDL, high-density lipoprotein; LDL, low-density lipoprotein; CRP, C reactive protein. ^*^HOMA-IR index calculated as fasting insulin (mU/l) × FBS (mmol/l)/22.5

**ESM Table 2**. Characteristics of the included participants for different type 2 diabetes sub-phenotypes in the PROSPER Study

| **T2D Sub-phenotypes** | | **Dyslipidemic**  **sub-phenotype** | **Unspecified**  **sub-phenotype** | **Insulin-resistant**  **sub-phenotype** | ***P-value*** |
| --- | --- | --- | --- | --- | --- |
| No. of participants | | 46 | 93 | 40 |  |
| Age, years | | 75.10 (3.13) | 75.09 (3.06) | 75.46 (3.23) | 0.805 |
| Sex, male, % | | 19 (41.3) | 39 (41.9) | 30 (75.0) | 0.001 |
| Body mass index, kg/m^2^ | | 24.99 (3.24) | 27.97 (3.62) | 31.39 (4.12) | <0.001 |
| Fasting blood glucose, mmol/L | | 5.96 (1.16) | 8.50 (2.48) | 8.28 (2.34) | <0.001 |
| HOMA.IR^*^, median (IQR) | | 1.95 [1.40, 2.31] | 3.75 [2.87, 5.00] | 8.67 [6.58, 11.25] | <0.001 |
| Current smoking, % | | 15 (32.6) | 14 (15.1) | 6 (15.0) | 0.035 |
| Use of lipid-lowering medication, % | | 0 | 0 | 0 | NA |
| Use of blood pressure-lowering medication, % | | 25 (54.3) | 59 (63.4) | 36 (90.0) | 0.001 |
| Systolic blood pressure, mmHg | | 172.72 (21.59) | 176.46 (24.65) | 169.28 (20.44) | 0.238 |
| Total cholesterol, mg/L | | 224.33 (36.57) | 215.51 (26.90) | 199.85 (29.00) | 0.001 |
| HDL, mg/L | | 48.17 (12.95) | 43.87 (8.51) | 41.16 (9.51) | 0.005 |
| LDL, mg/L | | 150.43 (32.87) | 146.12 (21.90) | 129.12 (28.49) | 0.001 |
| CRP, mg/L | | 2.60 [1.65, 6.56] | 3.39 [2.06, 6.51] | 4.99 [2.87, 7.74] | 0.034 |
| Polygenic risk score | | 276.16 [270.70, 285.95] | 280.31 [270.35, 287.46] | 280.87 [271.89, 287.72] | 0.456 |
| Tertile of polygenic risk score, % | |  |  |  |  |
|  | Lower | 16 (34.8) | 32 (34.4) | 12 (30.0) | 0.951 |
|  | Medium | 16 (34.8) | 29 (31.2) | 15 (37.5) |  |
|  | High | 14 (30.4) | 32 (34.4) | 13 (32.5) |  |

**Abbreviations:** SD, standard deviation; HOMA-IR, Homeostatic Model Assessment of Insulin Resistance; IQR, interquartile range; HDL, high-density lipoprotein; LDL, low-density lipoprotein; CRP, c-reactive protein. ^*^HOMA-IR index calculated as fasting insulin (mU/l) × FBS (mmol/l)/22.5

**ESM Figure 1**. BIC value for the cluster analysis

**Rotterdam Study**

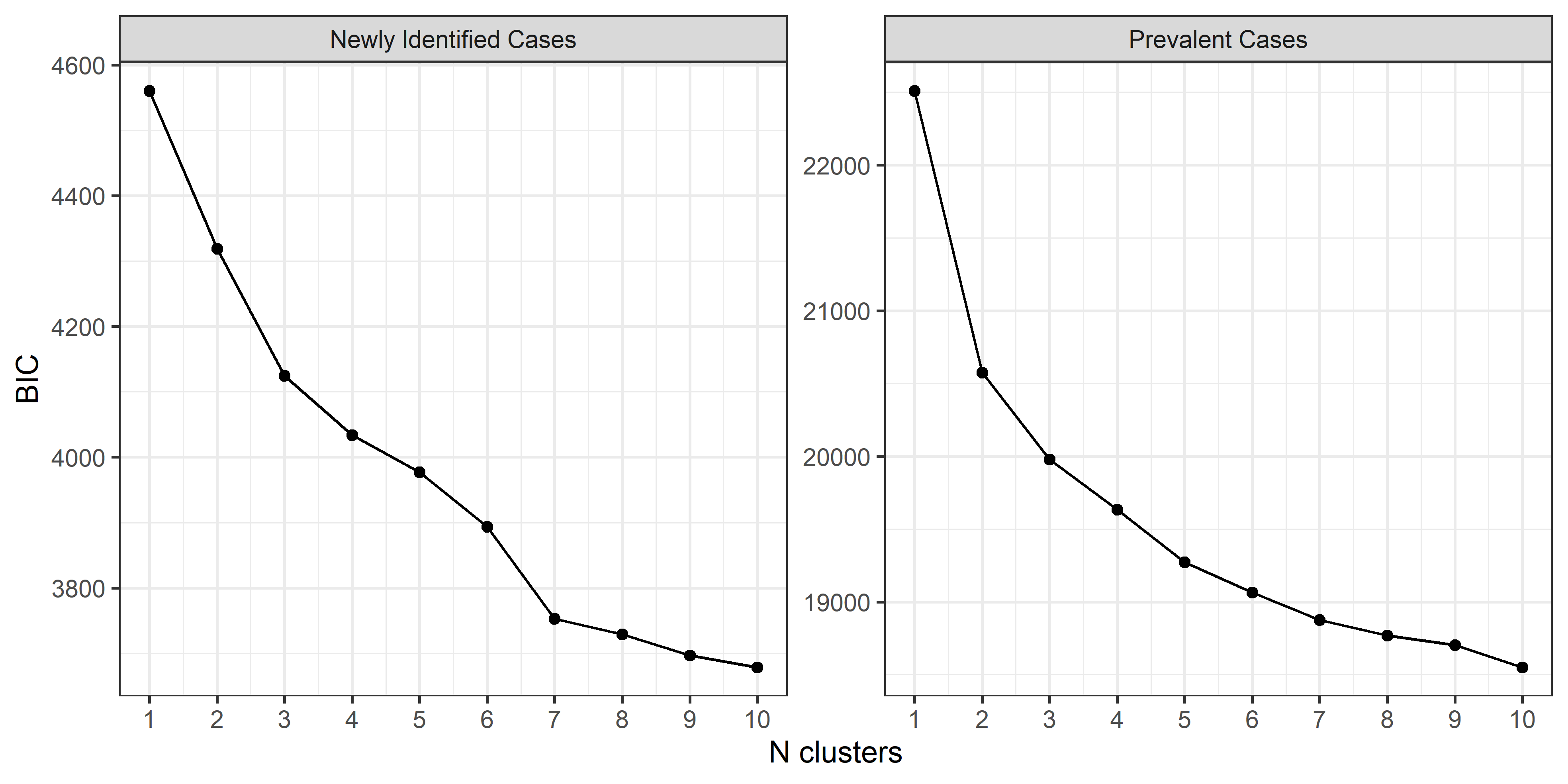

**KORA Study**

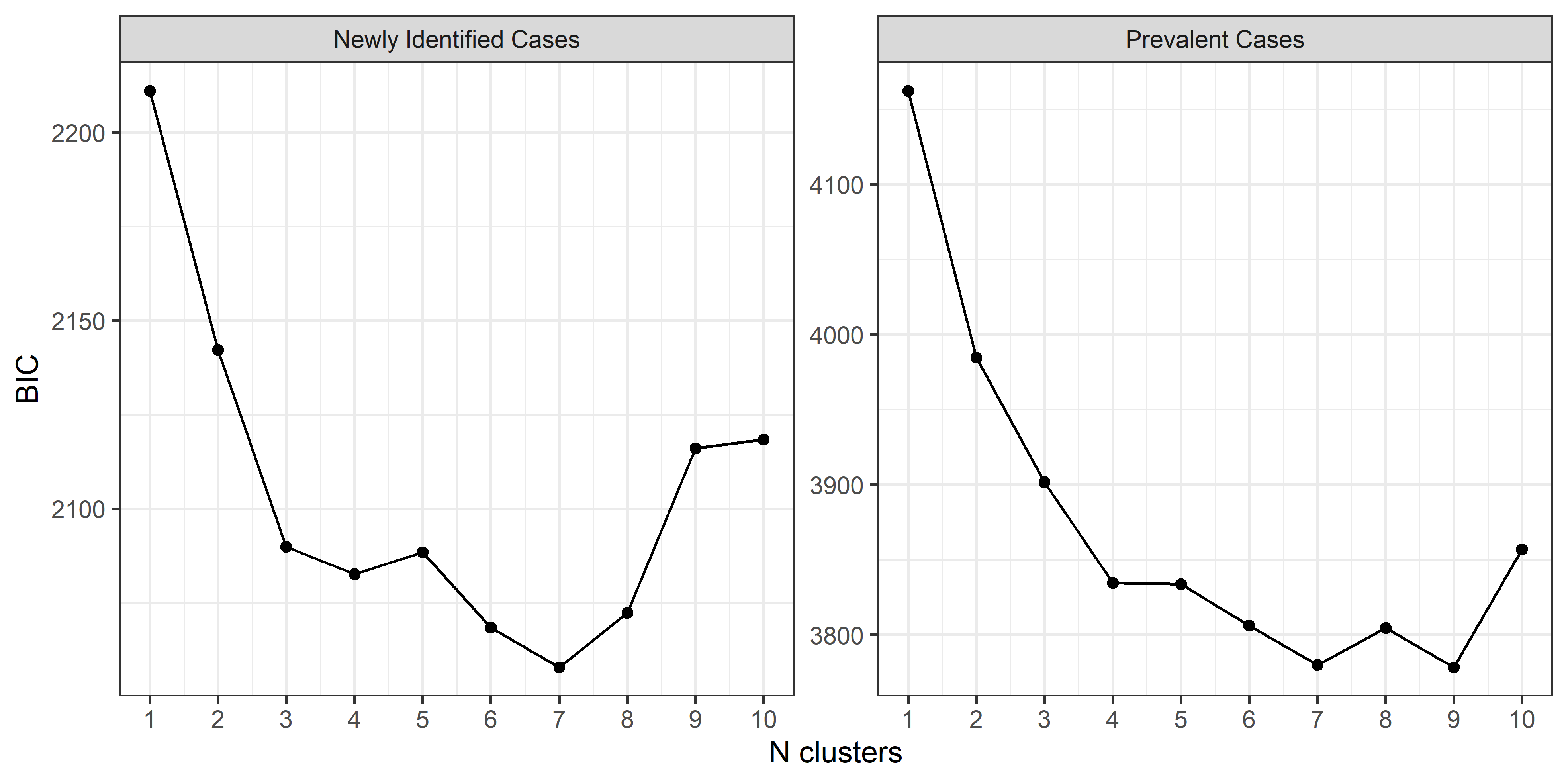

**PROSPER Study**

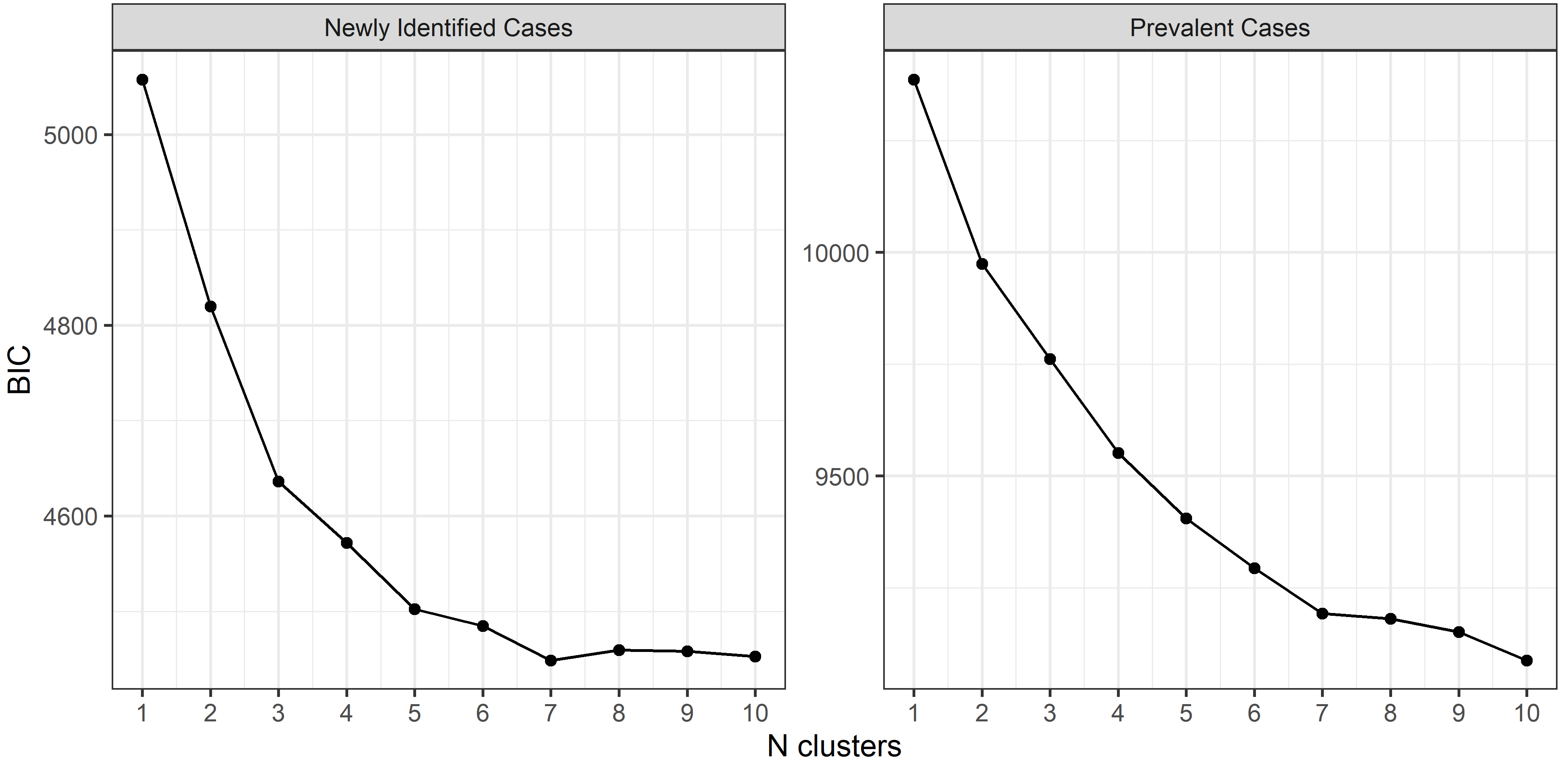

**ESM Figure 2**. Average silhouette for the cluster analysis

**Rotterdam Study**

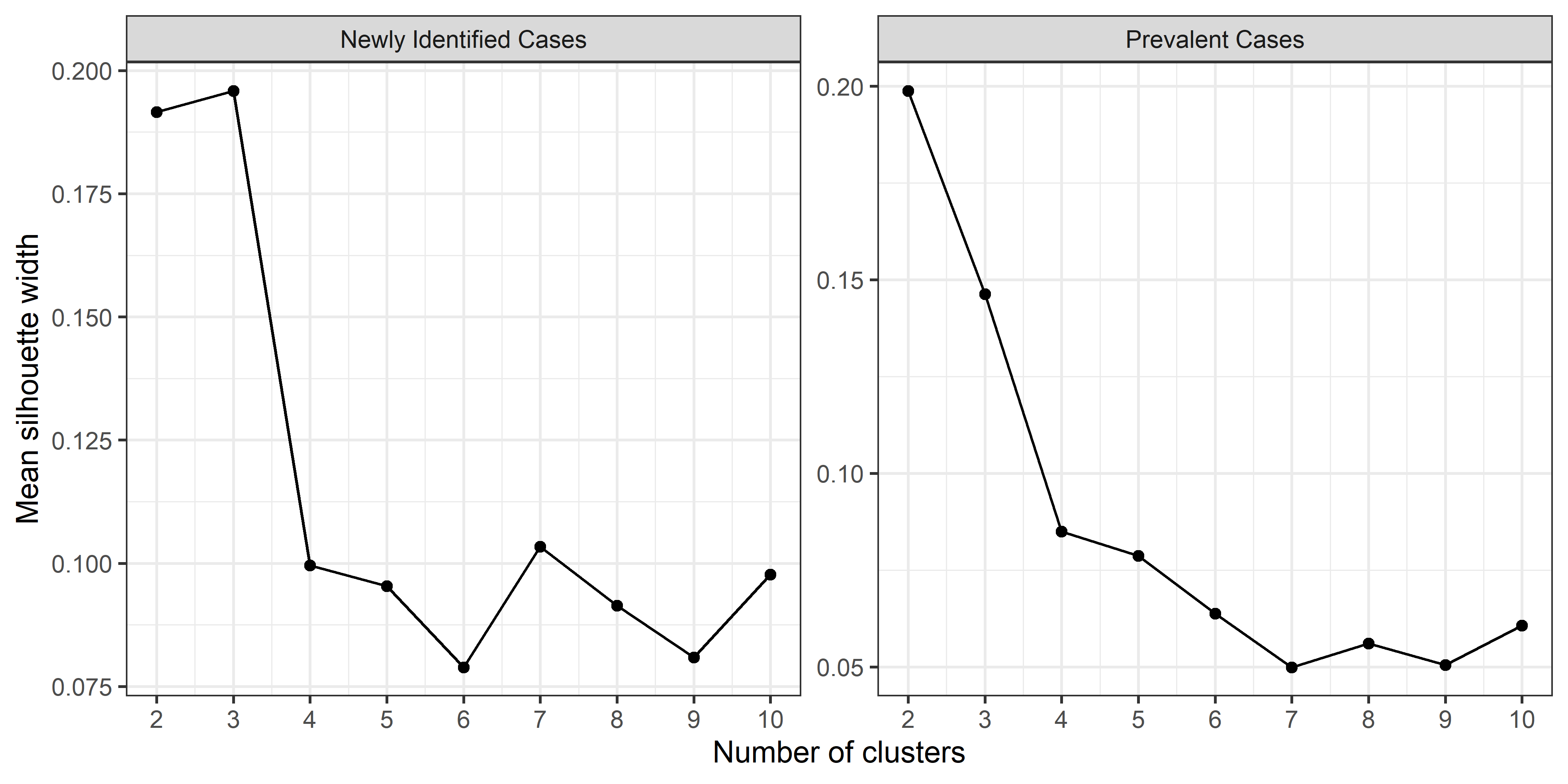

**KORA Study**

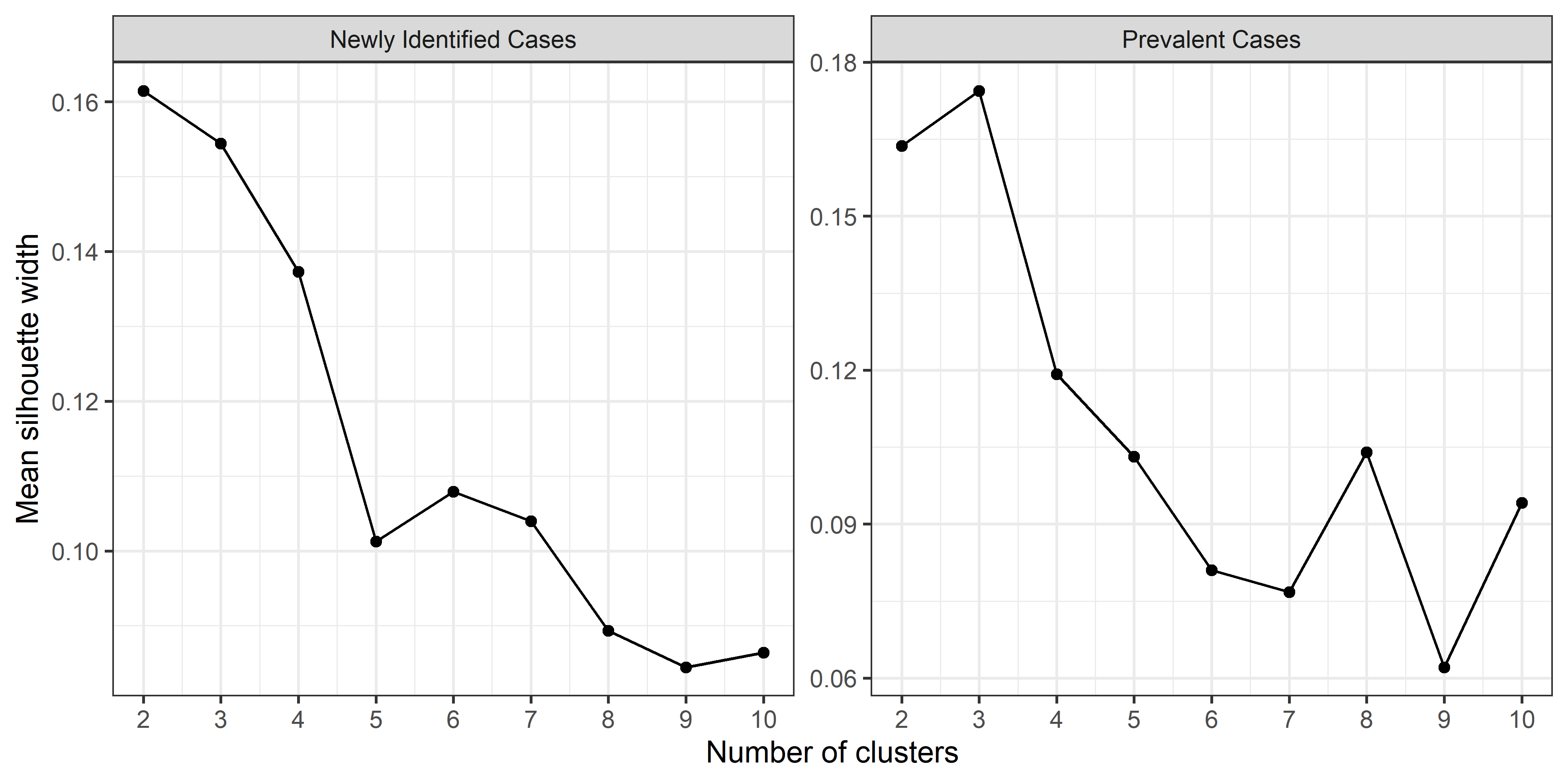

**PROSPER Study**

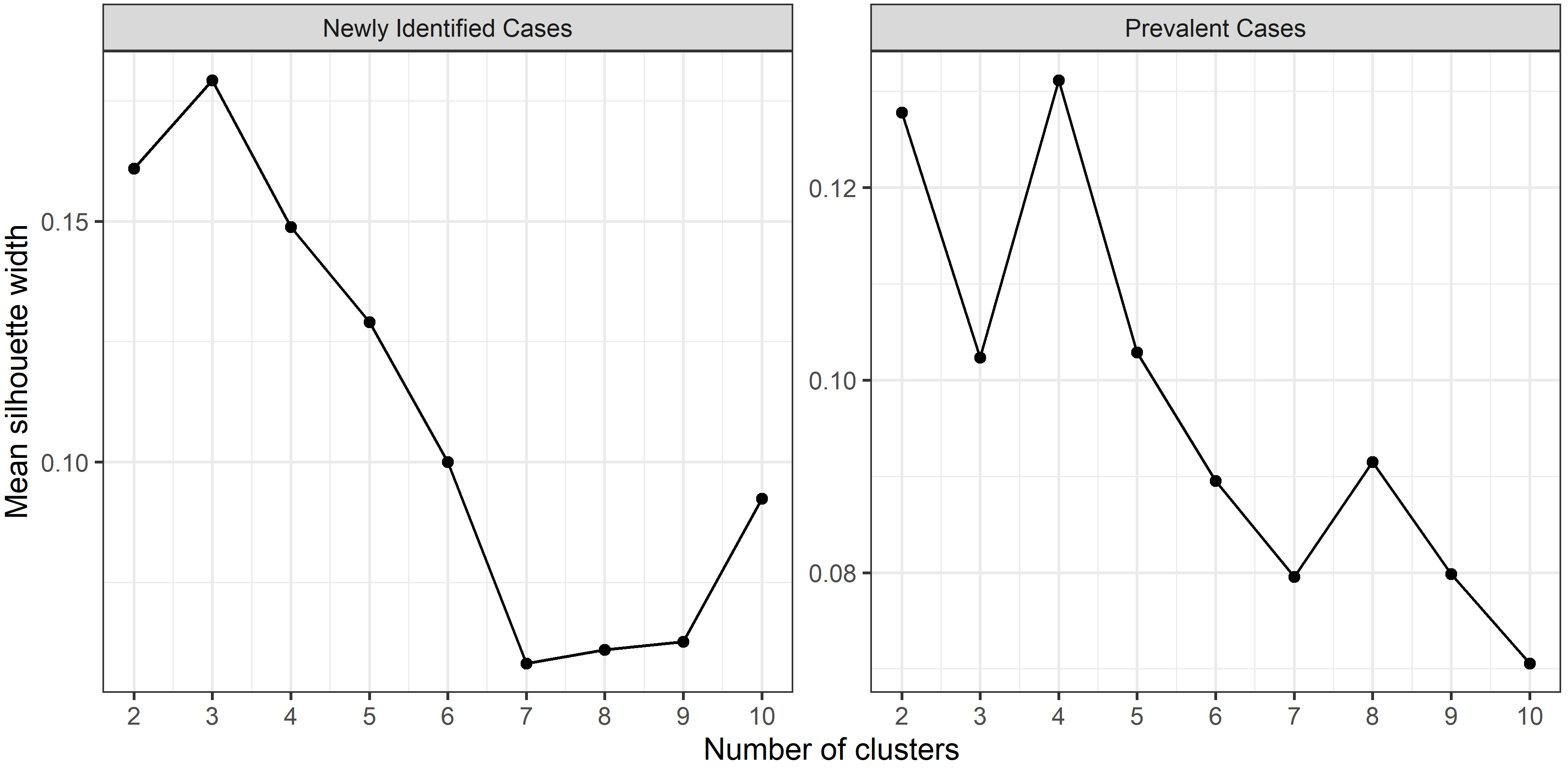

**ESM Figure 3**. Gap statistic for the cluster analysis

**Rotterdam Study**

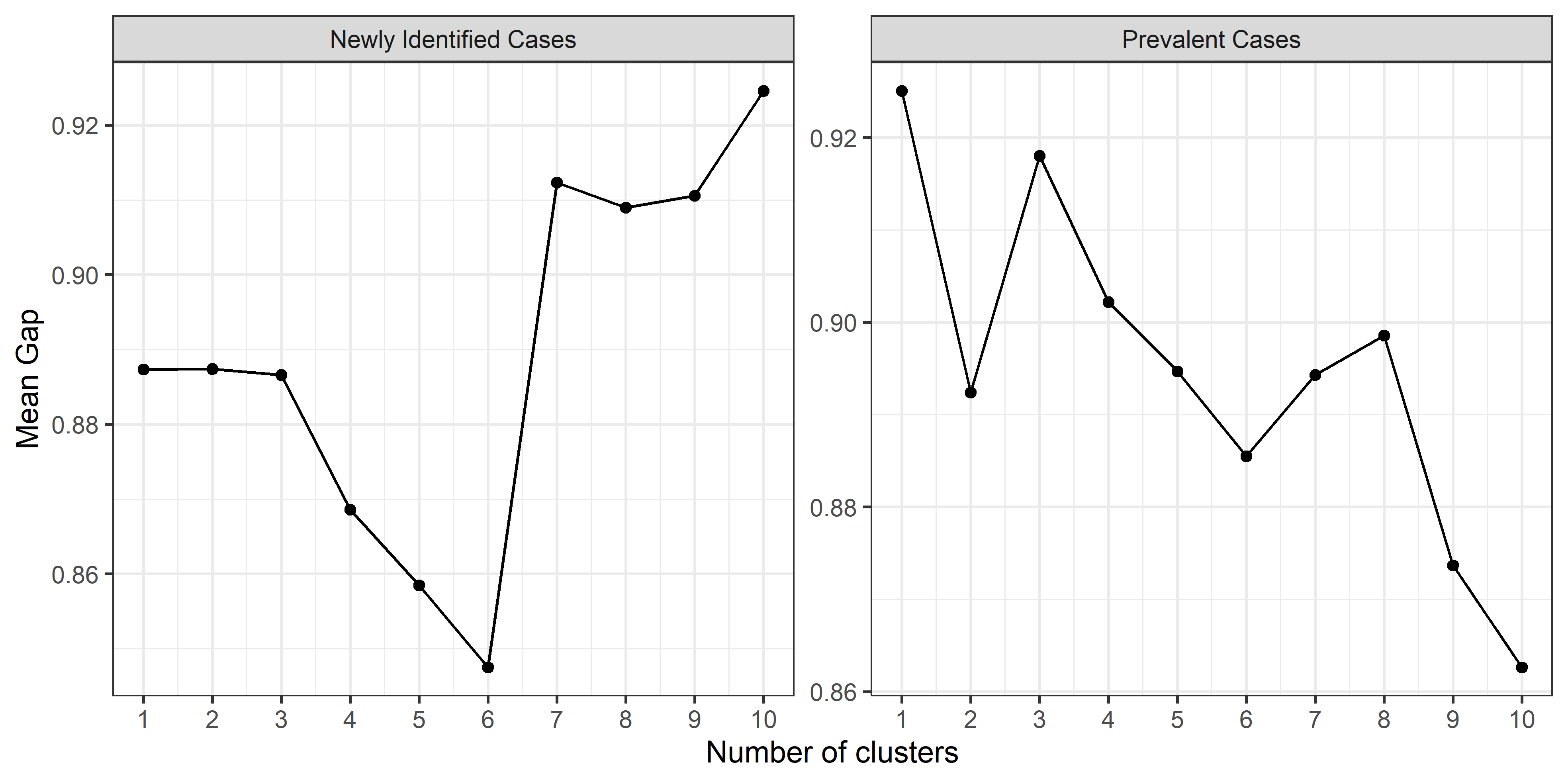

**KORA Study**

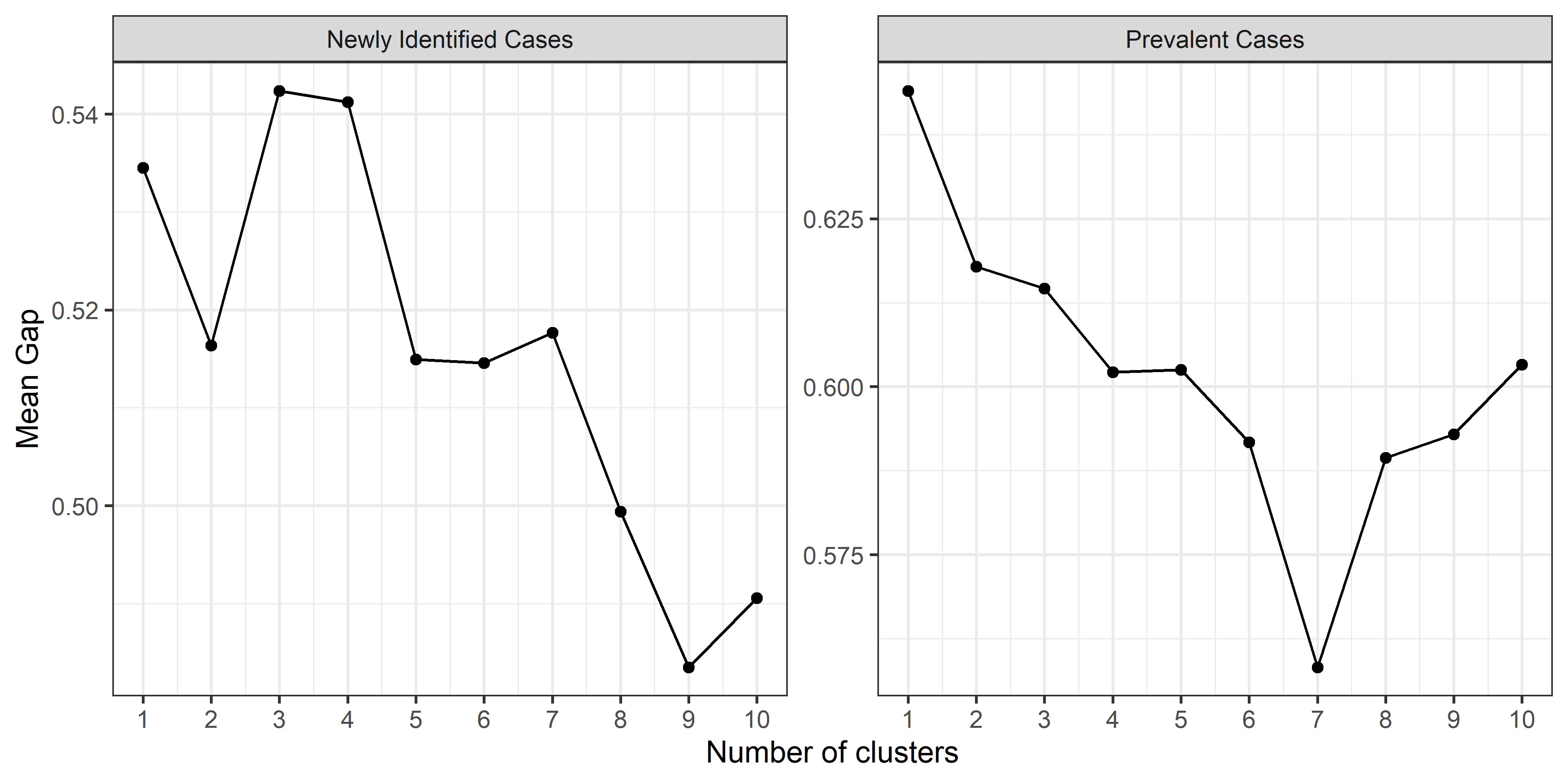

**PROSPER Study**

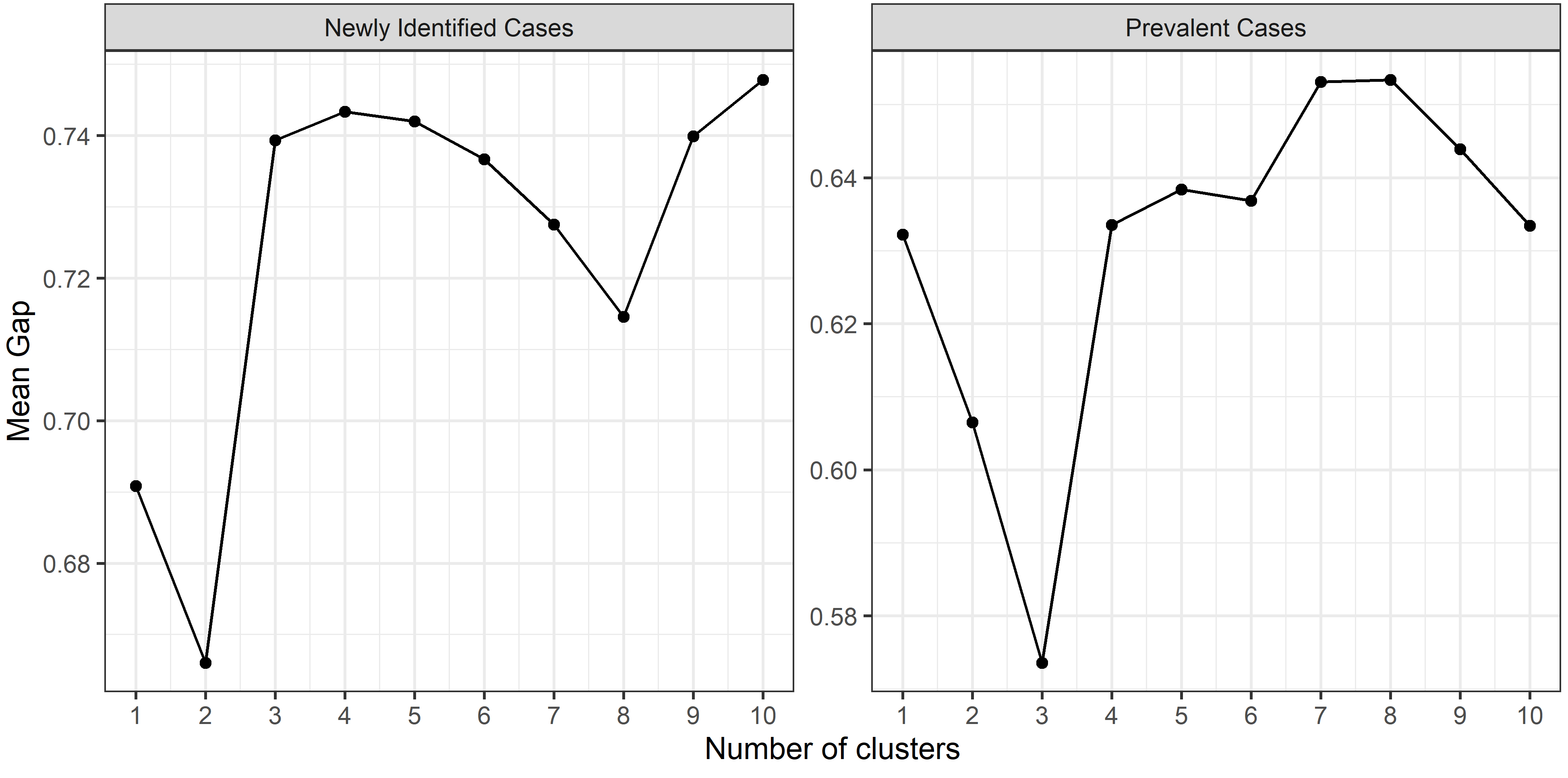

**ESM Figure 4**. Survival plot of incident cardiovascular disease among different type 2 diabetes sub-phenotypes

**Rotterdam Study**

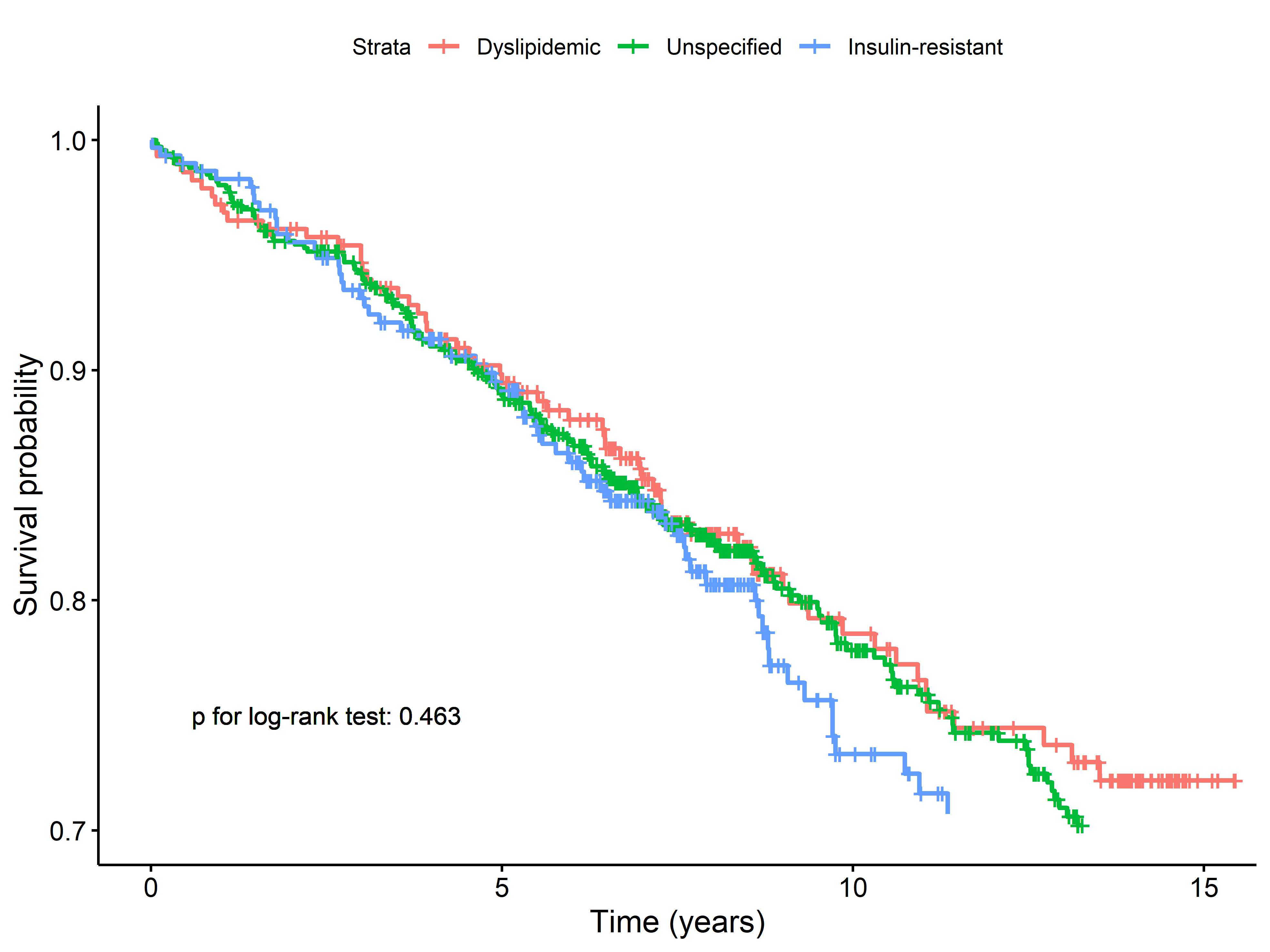

**KORA Study**

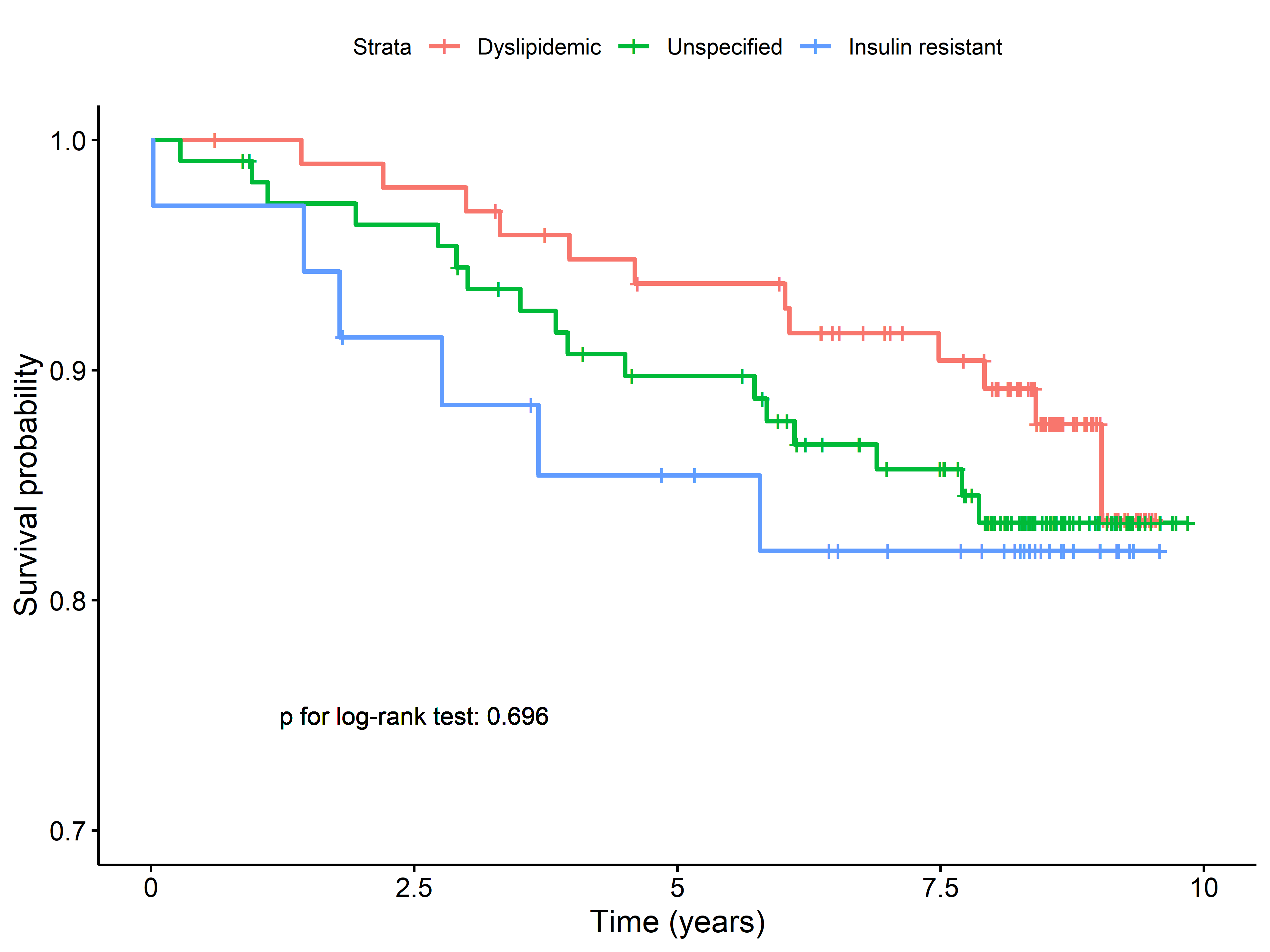
